# Artificial Intelligence-Based Detection of Airway Mucus Plugs on CT and Associations With Clinical Outcomes in COPDGene

**DOI:** 10.64898/2026.06.10.26355393

**Authors:** John Oyer, Ali Namvar, Benjamin A. Hoff, Christopher Bosma, Wassim W. Labaki, Ella A. Kazerooni, Fernando J. Martinez, Charles R. Hatt, MeiLan K. Han, Craig J Galban, Sundaresh Ram

## Abstract

**RATIONALE:** Airway mucus plugging is a clinically relevant manifestation of airway pathology in chronic obstructive pulmonary disease (COPD) and is associated with increased mortality even in early disease; however, visual computed tomography (CT) assessment is subjective and labor intensive.

**OBJECTIVES:** To develop an AI–based quantitative CT method for automated detection of airway mucus plugging and evaluate associations with physiologic impairment and clinical outcomes.

**METHODS:** Inspiratory CT scans from 8,971 COPDGene Phase 1 (GOLD 0–4 and PRISm) participants were analyzed. An AI-based framework combining 3D airway segmentation discontinuities and convolutional neural network classification identified mucus plug obstructions, yielding mucus plug burden (total plug count). Associations with outcomes were evaluated using covariate-adjusted models.

**MEASUREMENTS AND MAIN RESULTS:** Higher mucus plug burden was associated with lower post-bronchodilator FEV₁ % predicted (ρ = −0.41; P < 0.001), greater air trapping (LAA < −856 HU; ρ = 0.33; P < 0.001), worse health status (SGRQ; ρ = 0.31; P < 0.001), and shorter 6-minute walk distance (ρ = −0.26; P < 0.001). Among GOLD 1–4 participants, mucus plug presence was independently associated with increased all-cause mortality (adjusted hazard ratio, 1.28; P < 0.005) and exacerbation frequency (adjusted incidence rate ratio, 1.32; P < 0.005). Plug presence was also associated with increased respiratory mortality across GOLD categories and cardiovascular mortality in GOLD 1–2.

**CONCLUSIONS:** AI-based quantitative CT assessment of airway mucus plugging provides a scalable, reproducible measure associated with physiologic impairment and adverse outcomes in COPD, supporting its role in risk stratification and future therapeutic studies.

## Introduction

Airway mucus plugging is a characteristic manifestation of muco-obstructive airway disease observed across multiple chronic lung disorders, including chronic obstructive pulmonary disease (COPD), asthma, cystic fibrosis, bronchiectasis, and primary ciliary dyskinesia (1–6). Across these conditions, abnormal mucus production, impaired mucociliary clearance, and altered mucus rheology promote intraluminal secretion retention, resulting in partial or complete airway obstruction. Imaging and pathologic studies have established mucus plugging as a structural correlate of airway dysfunction that is detectable on computed tomography (CT) and relevant to disease expression across a range of airway-centered phenotypes.

COPD is a major global cause of morbidity and mortality and remains among the leading causes of death worldwide (7, 8). It is clinically heterogeneous, with wide variation in symptoms, exacerbation risk, and outcomes that are incompletely captured by spirometric severity alone (9, 10). Within this heterogeneity, CT-visible airway mucus plugging has emerged as a clinically informative airway phenotype associated with physiologic impairment and adverse outcomes independent of emphysema and other CT measures of airway disease (11–14). However, these observations have largely relied on visual CT scoring approaches that are time-consuming, reader dependent, and difficult to scale for large, with substantial interobserver variability. Existing automated methods remain limited and often depend on plug-level manual annotation or synthetic training strategies (15), constraining reproducibility and broader applicability. As a result, scalable and objective quantification of mucus plugging remains a key unmet need for large cohort studies and clinical translation.

We therefore developed an automated, artificial intelligence (AI)-based quantitative CT framework for airway mucus plug detection that leverages an anatomic premise: focal airway occlusion manifests as discontinuities in airway segmentations. This approach integrates three-dimensional airway segmentation with a convolutional neural network based-classification to identify mucus plug-associated quantify plug burden without requiring manual plug annotations. We hypothesized that automated AI-based quantification of airway mucus plugging would reproducibly identify mucus plug burden and recapitulate its established associations with physiologic impairment and adverse clinical outcomes while enabling scalable and objective phenotyping. We applied this framework to inspiratory CT scans from nearly 9,000 participants obtained at the baseline (Phase 1) COPDGene study visit and evaluated associations with lung function, CT-based air trapping, symptom burden, exercise capacity, exacerbations, and mortality. Preliminary findings were presented in abstract form at the American Thoracic Society International Conference; here we report comprehensive analyses and expanded evaluation of clinical outcomes.

## Methods

### Study population

The NIH-funded Genetic Epidemiology of Chronic Obstructive Pulmonary Disease (COPDGene®) study is a large, multicenter observational cohort that enrolled participants aged 45–80 years with a smoking history of at least 10 pack-years, including individuals with and without airflow obstruction (16). All participants underwent extensive baseline characterization, including standardized symptom assessment, pre- and post-bronchodilator spirometry, and quantitative computed tomography (CT) imaging, as previously described (16). COPD was defined by a post-bronchodilator FEV₁/FVC ratio <0.70 in accordance with Global Initiative for Chronic Obstructive Lung Disease (GOLD) criteria (17).

For the present analysis, we included 8,971 participants from Phase 1 (baseline visit) of COPDGene with available inspiratory CT scans. The study population encompassed individuals across GOLD stages 1–4, as well as individuals classified as GOLD 0 (defined as normal spirometry regardless of the presence of conditions) and those with preserved ratio impaired spirometry (PRISm). Inspiratory CT images acquired at total lung capacity were used for automated detection and quantification of airway mucus plugging. Mucus plug burden was evaluated in relation to lung function, quantitative CT measures, symptoms, exercise capacity, exacerbations, and mortality.

### Automated Mucus Plug Detection

Automated mucus plug detection was performed using a three-step framework consisting of airway segmentation, airway discontinuity identification, and obstruction classification (**Fig 1**).

**Figure 1.**
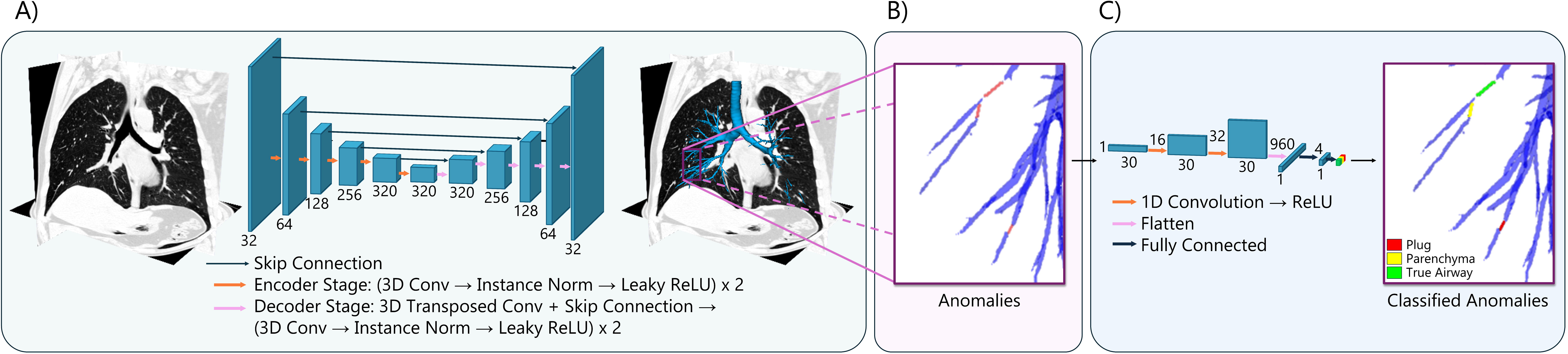
Automated mucus plug detection and classification workflow. (A) Inspiratory CT scans are processed using a 3D U-Net architecture with encoder–decoder stages and skip connections to generate a segmented airway tree. (B) Potential anomalies along the airways are extracted for further analysis. (C) A 3D convolutional neural network classifies each anomaly into mucus plug, airway, or parenchyma.

First, airway segmentation was performed using a deep learning model based on the nnU-Net architecture (18), which has demonstrated robust performance for medical image segmentation. The model was trained on inspiratory CT scans from the ATM’22 dataset (19), which includes high-quality scans with manual airway lumen annotations provided by three expert radiologists. The dataset was split into 209 scans for training and 70 scans for testing. Importantly, training annotations included only the airway lumen and excluded the airway wall, resulting in a model optimized to segment unobstructed, healthy airways.

Second, airway discontinuities were identified in the segmented COPDGene airway trees using a parametric graph-based airway continuity algorithm designed to detect anatomically plausible breaks in the airway tree structure. Candidate discontinuities were defined as disconnected airway components and were evaluated using geometric criteria based on endpoint connectivity, airway branch direction, and connection distance to distinguish likely luminal obstruction from segmentation fragmentation. Candidate discontinuities satisfying predefined plausibility criteria were retained as regions for downstream classification, restricting subsequent analysis to localized airway segments with potential mucus obstruction. (See **Supplemental Methods I. A** for the details).

Third, each candidate discontinuity was classified using a lightweight convolutional neural network (CNN) applied to one-dimensional voxel patches sampled along the airway centerline spanning the break and adjacent airway segments. Training data for unobstructed airway and parenchyma classes were sampled from visually confirmed regions. For the Mucus plug class, training examples were generated using a weakly supervised model-based strategy in which attenuation profiles from a limited number of visually confirmed complete occlusions were modeled using kernel density estimation with an Epanechnikov kernel, enabling representative plug modeling without extensive plug-level annotation (See **Supplemental Methods I. B** for the details). The trained classifier was then applied to all candidate discontinuities to quantify mucus plug burden. This trained model was then applied to inspiratory CT scans from all Phase 1 COPDGene participants with available imaging. Additional details regarding the nnU-Net airway segmentation model and the CNN classifier model training and implementation are provided in **Supplemental Methods I. C**.

Three measures of mucus plug burden were evaluated: mucus plug count, affected airways score, and Weibel sum (see **Supplemental Methods Section I. D** for definitions). Associations with physiologic and clinical outcomes were similar across all three measures (**Supplemental Figure 4**). Because mucus plug count performed comparably while remaining the simplest and most interpretable metric, it was selected as the primary measure of mucus plug burden. Accordingly, mucus plug burden refers to total mucus plug count throughout the remainder of the manuscript unless otherwise specified.

### Statistical Analysis

Associations between mucus plug burden (total plug count) and outcomes were evaluated using regression models. Spearman’s rank correlation assessed unadjusted associations between mucus plug count and continuous measures (post-bronchodilator FEV₁ % predicted, PRM air trapping, SGRQ, and 6-minute walk distance). PRM air trapping (%) was calculated as the sum of PRM emphysema (%) and PRM-defined fSAD (%).

All-cause mortality was analyzed with multivariable Cox proportional hazards models, with Kaplan–Meier curves stratified by mucus plug count. Cause-specific mortality was analyzed using Fine–Gray sub distribution hazards models to account for competing risks (20). Retrospectively reported exacerbations were defined as episodes requiring treatment with antibiotics and/or systemic corticosteroids. Severe exacerbations were defined as exacerbations requiring hospitalization. Presence of severe exacerbations was analyzed with logistic regression, and exacerbation frequency with negative binomial regression.

For Cox, logistic, and negative binomial models, two covariate sets were used. Model 1 included age, sex, race, pack-years, current smoking status, FEV₁ % predicted, CT emphysema (% voxels ≤ −950 HU), and Pi10 (standardized airway wall thickness). Model 2 included Model 1 covariates plus self-reported asthma, chronic bronchitis, coronary artery disease, 6-minute walk distance, and BODE index; Cox Model 2 additionally included exacerbation frequency and history of severe exacerbations. Fine–Gray models included Model 1 covariates plus coronary artery disease, congestive heart failure, stroke, diabetes, hypertension, and lung, breast, prostate, colon, and bladder cancer.

Emphysema (%) and Pi10 were derived using Thirona software (Nijmegen, the Netherlands), and PRM using Lung Density Analysis software (Imbio LLC, Minneapolis, MN, USA). Analyses were performed in Python with a two-sided significance level of 0.05.

## Results

Baseline characteristics and clinical outcomes, overall and stratified by mucus plug burden, are summarized in **Table 1**. Alternative burden metrics incorporating airway obstruction extent and airway-generation weighting produced findings similar to those obtained using mucus plug count (**Supplemental Figure 4**), supporting the use of total plug count as the primary burden measure in subsequent analyses.

**Table 1.** Detected mucus plug features from COPDGene Phase 1.

| Category | All | 0 plugs | 1 plug | 2 plugs | 3 plugs | 4 plugs | ≥5 plugs |
| --- | --- | --- | --- | --- | --- | --- | --- |
| **Age (y), mean (SD) [No.]** | 59.6 (9.0)<br>[8971] | 58.2 (8.6)<br>[5646] | 60.7 (9.4)<br>[1634] | 62.0 (9.1)<br>[638] | 61.9 (9.1)<br>[348] | 64.0 (8.8)<br>[222] | 64.0 (9.0)<br>[483] |
| **Sex** |  |  |  |  |  |  |  |
| **Male** | 4803 (53.5%) | 3060 (54.2%) | 792 (48.5%) | 324 (50.8%) | 194 (55.7%) | 126 (56.8%) | 307 (63.6%) |
| **Female** | 4168 (46.5%) | 2586 (45.8%) | 842 (51.5%) | 314 (49.2%) | 154 (44.3%) | 96 (43.2%) | 176 (36.4%) |
| **Race** |  |  |  |  |  |  |  |
| **Non-Hispanic White** | 6087 (67.9%) | 3790 (67.1%) | 1097 (67.1%) | 413 (64.7%) | 242 (69.5%) | 165 (74.3%) | 380 (78.7%) |
| **Non-Hispanic Black** | 2884 (32.1%) | 1856 (32.9%) | 537 (32.9%) | 225 (35.3%) | 106 (30.5%) | 57 (25.7%) | 103 (21.3%) |
| **BMI, mean (SD) [No.]** | 28.8 (6.2)<br>[8971] | 29.1 (6.1)<br>[5646] | 28.9 (6.5)<br>[1634] | 28.5 (6.5)<br>[638] | 27.9 (6.2)<br>[348] | 28.2 (6.4)<br>[222] | 26.8 (6.0)<br>[483] |
| **Smoking status** |  |  |  |  |  |  |  |
| **Former smoker** | 4242 (47.3%) | 2555 (45.3%) | 805 (49.3%) | 328 (51.4%) | 175 (50.3%) | 135 (60.8%) | 244 (50.5%) |
| **Current smoker** | 4729 (52.7%) | 3091 (54.7%) | 829 (50.7%) | 310 (48.6%) | 173 (49.7%) | 87 (39.2%) | 239 (49.5%) |
| **Pack-years of smoking, mean (SD) [No.]** | 44.3 (24.9)<br>[8969] | 41.5 (22.8)<br>[5645] | 46.5 (25.8)<br>[1633] | 49.5 (29.0)<br>[638] | 51.4 (27.3)<br>[348] | 53.5 (27.1)<br>[222] | 53.7 (29.9)<br>[483] |
| **Medical history** |  |  |  |  |  |  |  |
| **Asthma** | 1695 (18.9%) | 852 (15.1%) | 377 (23.1%) | 172 (27.0%) | 95 (27.3%) | 63 (28.4%) | 136 (28.2%) |
| **Coronary artery disease** | 594 (6.6%) | 336 (6.0%) | 113 (6.9%) | 56 (8.8%) | 24 (6.9%) | 26 (11.7%) | 39 (8.1%) |
| **Chronic bronchitis** | 1480 (16.5%) | 704 (12.5%) | 335 (20.5%) | 147 (23.0%) | 97 (27.9%) | 56 (25.2%) | 141 (29.2%) |
| **GOLD Spirometric stage** | 3869/699/1736/ | 3088/495/904/ | 520/118/412/ | 131/34/171/ | 58/18/89/ | 30/20/52/ | 42/14/108/ |
| **(GOLD 0/1/2/3/4/PRISm)** | 1042/530/1092 | 341/104/712 | 254/113/217 | 136/87/79 | 91/55/36 | 62/42/16 | 158/129/32 |
| **BODE index, mean (SD) [No.]** | 1.8 (2.2)<br>[8838] | 1.2 (1.8)<br>[5594] | 2.2 (2.3)<br>[1602] | 3.1 (2.6) [624] | 3.3 (2.6) [337] | 3.6 (2.6) [215] | 4.1 (2.7) [466] |
| **FEV<sub>1</sub> (L), mean (SD) [No.]** | 2.2 (0.9)<br>[8968] | 2.5 (0.8)<br>[5644] | 2.0 (0.8)<br>[1634] | 1.7 (0.8) [638] | 1.6 (0.8) [347] | 1.5 (0.8) [222] | 1.4 (0.8) [483] |
| **FEV<sub>1</sub> (% predicted), mean (SD) [No.]** | 76.3 (25.5)<br>[8968] | 84.0 (21.4)<br>[5644] | 70.5 (24.8)<br>[1634] | 61.5 (26.1)<br>[638] | 58.0 (25.4)<br>[347] | 56.0 (27.4)<br>[222] | 48.4 (24.7)<br>[483] |
| **Emphysema on CT (%), mean (SD) [No.]** | 6.3 (9.9)<br>[8959] | 4.5 (7.7)<br>[5634] | 7.2 (10.6)<br>[1634] | 10.0 (12.8)<br>[638] | 11.1 (12.7)<br>[348] | 12.5 (13.6)<br>[222] | 13.2 (13.4)<br>[483] |
| **Standardized airway wall thickness (Pi10), mean (SD) [No.]** | 2.4 (0.6)<br>[8959] | 2.2 (0.5)<br>[5634] | 2.5 (0.6)<br>[1634] | 2.7 (0.6) [638] | 2.8 (0.7) [348] | 2.8 (0.6) [222] | 3.0 (0.7) [483] |
| **6-minute walk distance (ft), mean (SD) [No.]** | 1361.5<br>(397.3)<br>[8852] | 1436.6 (376.7)<br>[5601] | 1292.1 (381.4)<br>[1606] | 1191.5 (403.3)<br>[625] | 1189.9 (392.2)<br>[338] | 1160.2 (414.3)<br>[215] | 1144.0 (415.1)<br>[467] |
| **Exacerbation frequency, mean (SD) [No.]** | 0.4 (0.9)<br>[8971] | 0.3 (0.8)<br>[5646] | 0.5 (1.0)<br>[1634] | 0.6 (1.1) [638] | 0.8 (1.3) [348] | 0.7 (1.2) [222] | 1.0 (1.5) [483] |

SD = Standard Deviation. No. = number of cases. GOLD = Global Initiative for Chronic Obstructive Lung Disease. FEV<sub>1</sub> = Forced expiratory volume in the first second of exhalation. PRISm = Preserved Ratio Impaired Spirometry.
\*Due to missing data, age, sex, race, BMI, smoking status, exacerbation frequency, race, asthma, coronary artery disease and chronic bronchitis n=8,971; pack-years of smoking n=8,969; FEV<sub>1</sub>, FEV<sub>1</sub>% predicted n=8,968; 6-minute walk distance n=8852; BODE index n=8838; emphysema on CT %, Pi10 n=8959

Performance of the nnU-Net airway segmentation model (Step 1 of the three-step workflow) was evaluated on the independent ATM’22 test set (n=70). The model achieved a mean Dice similarity coefficient of 0.93 ± 0.02, true positive rate of 0.93 ± 0.04, Jaccard index of 0.88 ± 0.04, false positive rate (×10⁻⁴) of 0.55 ± 0.41, Hausdorff distance of 42.75 ± 20.53, and airway tree detection rate of 0.91 ± 0.06, demonstrating robust lumen segmentation performance on unseen data. Application of the full three-step workflow enabled identification of focal airway discontinuities consistent with mucus plug obstruction. **Figure 2** shows a representative example of a mucus plug on CT displayed using multiplanar reconstruction.

**Figure 2.**
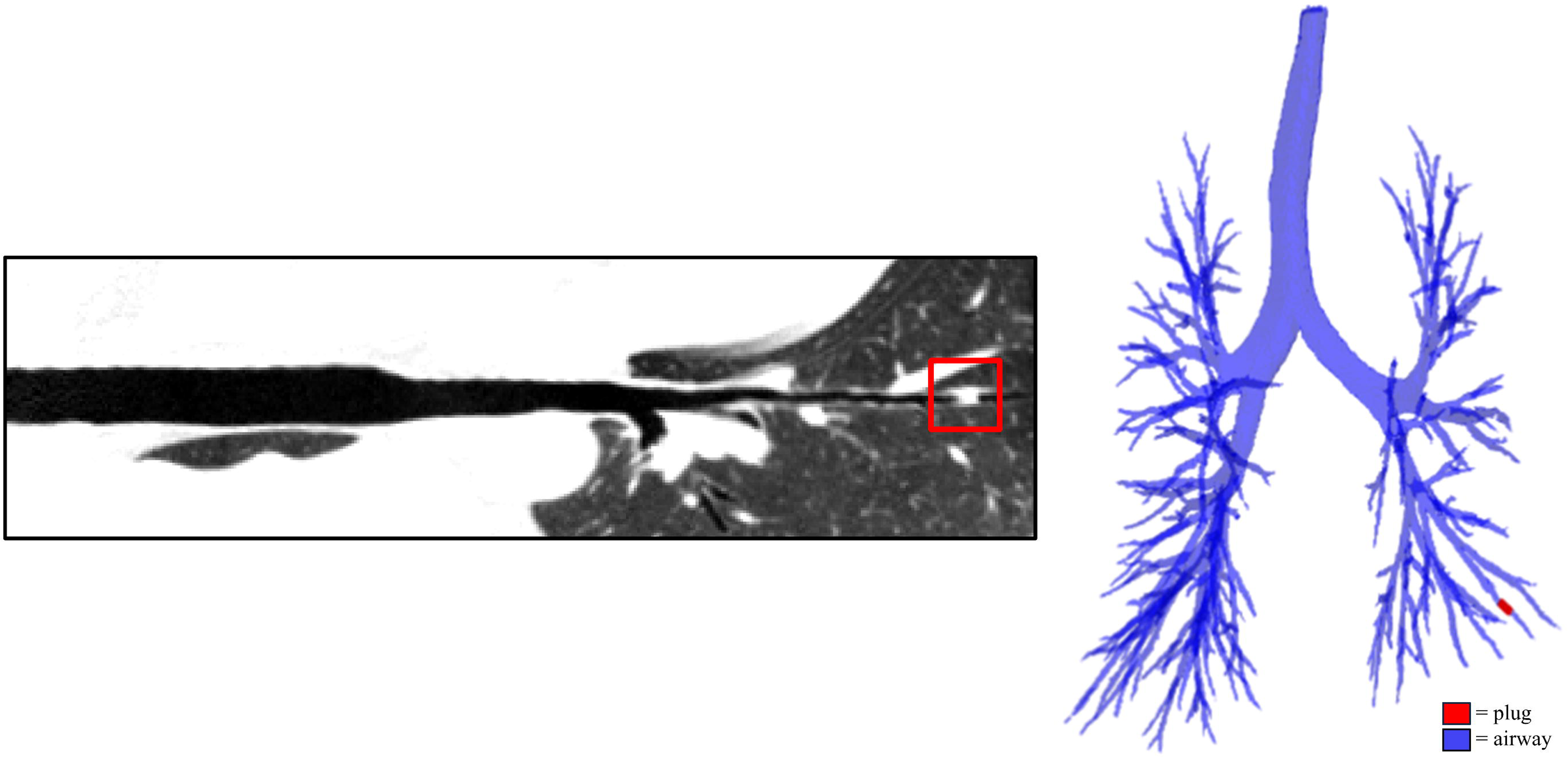
The leftmost figure shows a multiplanar reconstruction of a CT image demonstrating a mucus plug (red box) in an airway. The rightmost figure shows a 3D rendering of the complete airway tree with the corresponding plug highlighted in red. This case originates from the ATM ‘22 dataset and was not used for detection algorithm training.

Among 9,477 available Phase 1 COPDGene inspiratory CT scans, 8,971 participants had complete clinical, mortality, and imaging data and were included in the analysis (4,007 with airflow obstruction and 4,964 without).

### Association between mucus plugs and clinical/physiological outcomes

Mucus plug burden correlated inversely with post-bronchodilator FEV₁ % predicted (Spearman r = −0.41, P < 0.001; **Figure 3**) and positively with PRM air trapping (%) (Spearman r = 0.33, P < 0.001; Figure 1A in the online supplement). Higher plug burden was also associated with worse SGRQ scores (Spearman r = 0.31, P < 0.001; **Supplemental Figure 1C**) and shorter 6-minute walk distance (Spearman r = −0.26, P < 0.001; **Supplemental Figure 1B**). For display purposes, participants with ≥5 plugs were grouped; correlation analyses were conducted using continuous burden values. Mean mucus plug burden increased across GOLD stages (see **Supplemental Figure 2**).

**Figure 3.**
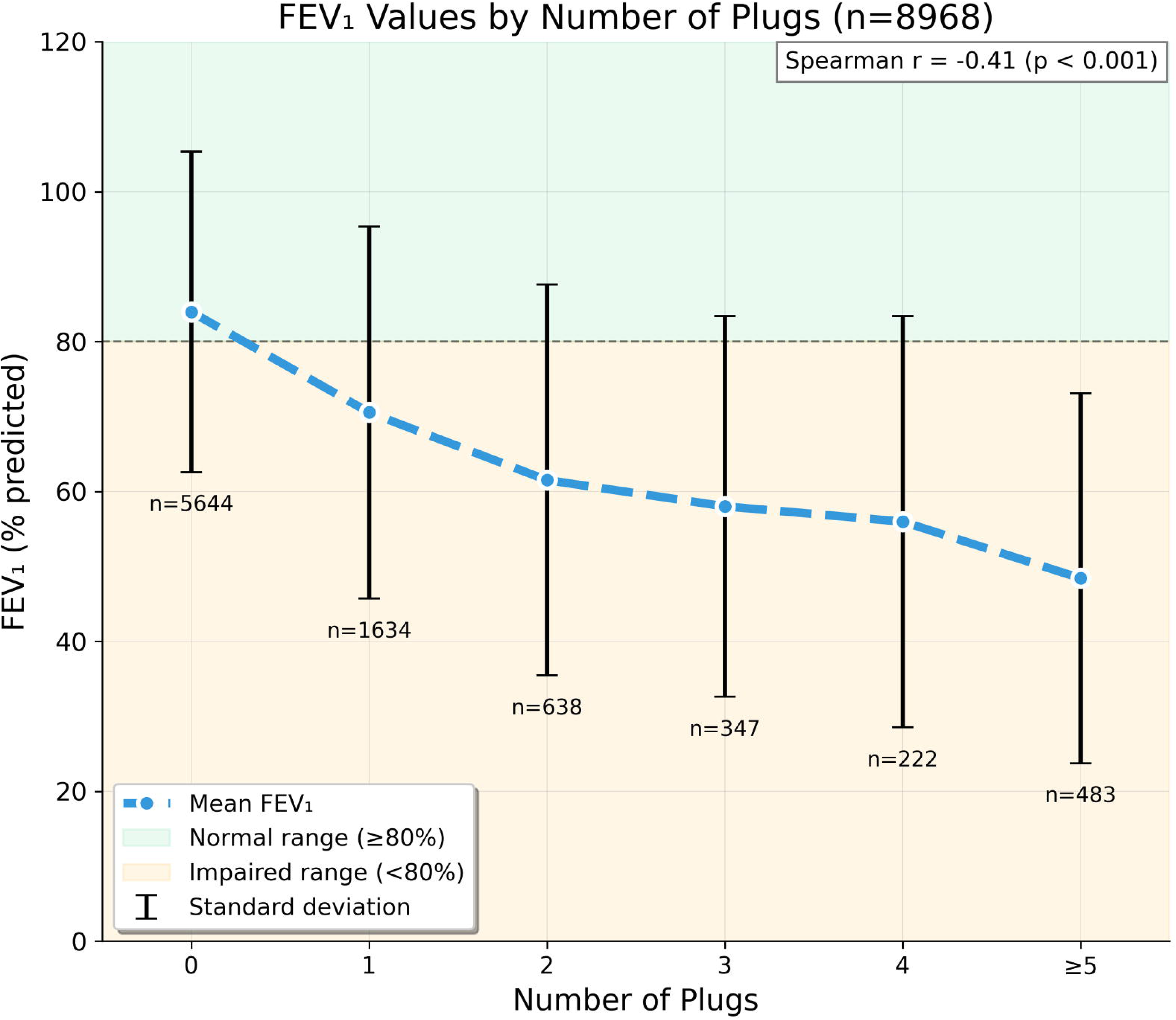
Relationship between the number of mucus plugs and FEV₁. The x-axis shows the number of plugs (0, 1, 2, 3, 4, ≥5), and the y-axis shows mean FEV₁ for each plug count, with error bars representing standard deviation. Shaded regions indicate the normal and impaired FEV₁ ranges. Spearman correlation coefficient (r) and p-value are displayed on the top right of the plot. For display purposes, participants with ≥5 plugs were grouped; correlation analysis was conducted using continuous burden values.

### Mucus plugs and exacerbations

Mucus plug presence was associated with higher exacerbation frequency and increased odds of severe exacerbations (**Table 2**). In unadjusted analyses, plug presence was associated with greater exacerbation frequency (rate ratio [RR], 1.74; P < 0.005) and severe exacerbations (odds ratio [OR], 2.07; P < 0.005). After adjustment, associations persisted for exacerbation frequency (adjusted rate ratio [aRR], 1.32 in Model 1 and 1.23 in Model 2; both P < 0.005) and severe exacerbations (adjusted odds ratio [aOR], 1.34 in Model 1 and 1.24 in Model 2; P < 0.005 and P = 0.03, respectively).

**Table 2.**
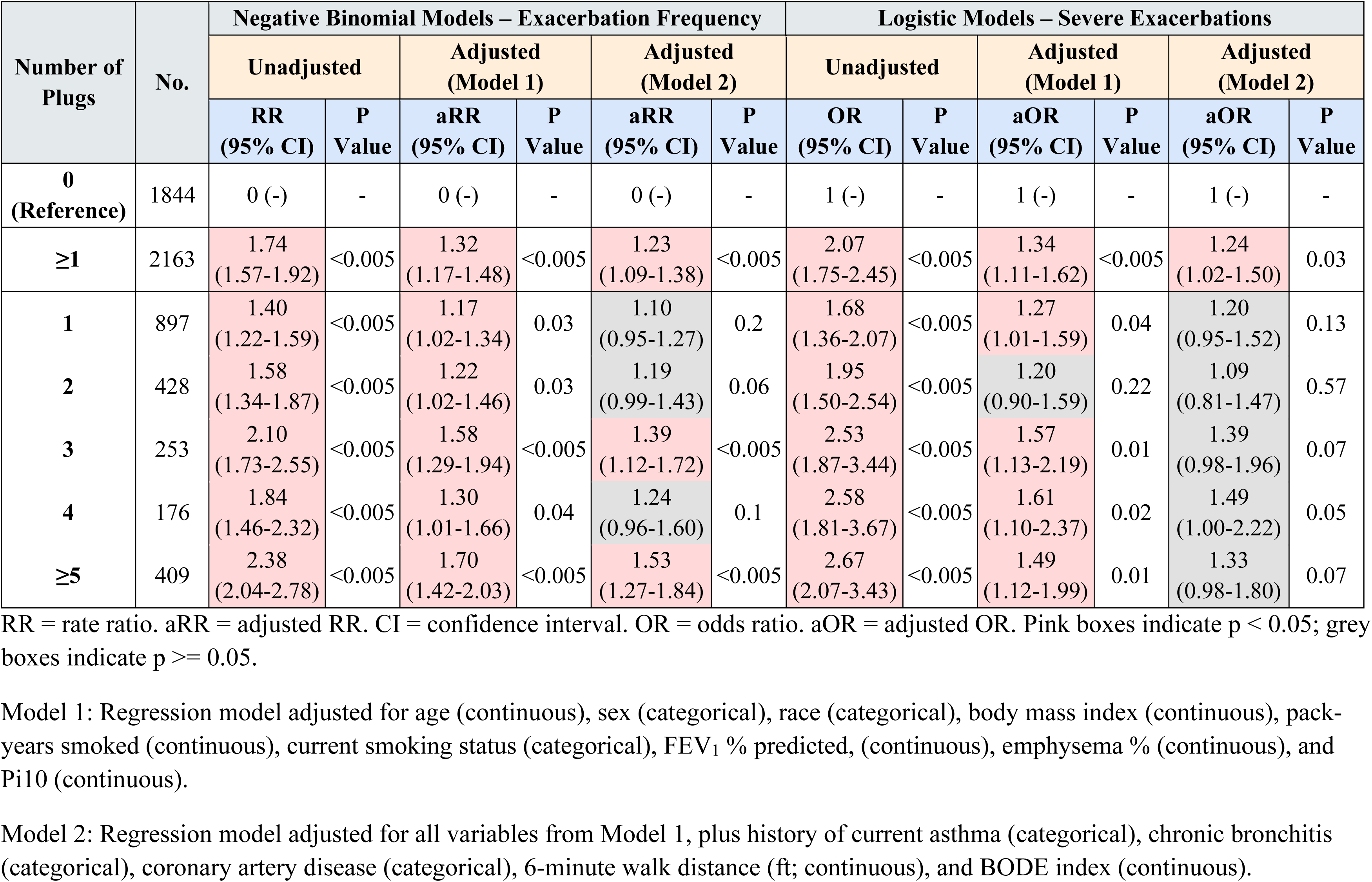
Results of exacerbation regression analysis for GOLD 1–4 cases.

### Mucus plugs and mortality

Among participants in GOLD stages 1–4, Kaplan–Meier analysis demonstrated progressively lower survival with increasing mucus plug burden over a median follow up of 8.9 years (IQR 4.2-12.1) (**Figure 4A**). Survival differed significantly across categories (0, 1, 2–4, and ≥5 plugs; global log-rank P < 0.0001). Because pairwise comparisons showed no significant differences among 2, 3, and 4 plugs, these categories were combined for primary visualization and analysis; separate curves are shown in **Supplemental Figure 2** in the online supplement.

**Figure 4.**
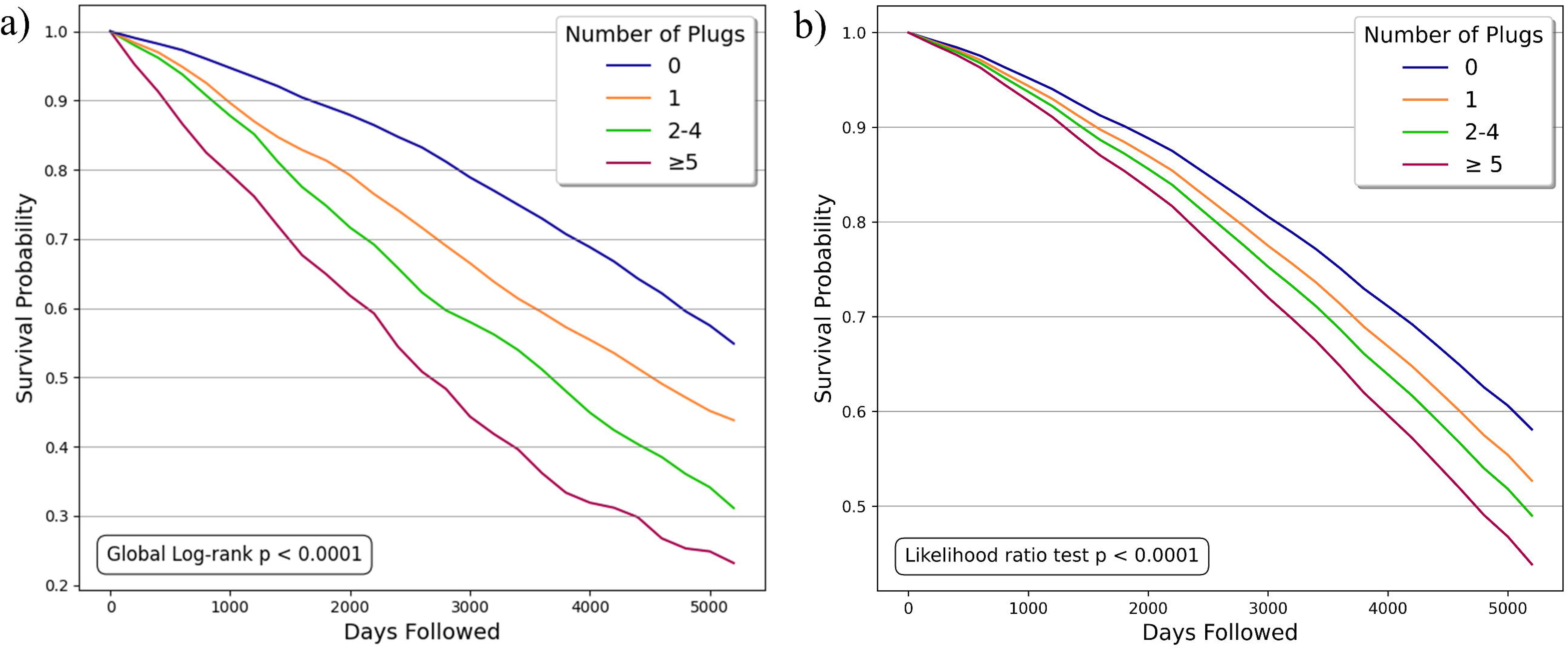
Results of survival analysis for GOLD 1–4 cases by number of plugs. Cases with 0, 1, 2–4, and ≥5 plugs at baseline are analyzed separately. Figure (a) shows unadjusted Kaplan–Meier analysis. Figure (b) shows survival probability predictions over time from an adjusted Cox proportional hazards model with adjustment for age, sex, race, body mass index, pack-years smoked, current smoking status, FEV_1_, emphysema %, and Pi10.

In Cox proportional hazards models, mucus plug burden was examined both as a binary variable (≥1 vs 0 plugs) and as a categorical variable (0, 1, 2, 3, 4, and ≥5 plugs) in relation to all-cause mortality (**Table 3**). In unadjusted analysis, the presence of any mucus plugs (≥1 vs 0) was associated with higher mortality risk (hazard ratio [HR], 1.96; 95% CI, 1.78–2.17; P < 0.005). The association remained significant after adjustment for demographics, smoking, lung function, and CT measures (Model 1 adjusted HR [aHR], 1.28; 95% CI, 1.15–1.42; P < 0.005) and persisted after further adjustment for comorbidities, functional measures, and exacerbation history (Model 2 aHR, 1.21; 95% CI, 1.09–1.35; P < 0.005). Model 1–derived survival curves are shown in **Figure 4B**, with significant differences across plug categories (likelihood ratio P < 0.0001).

**Table 3.**
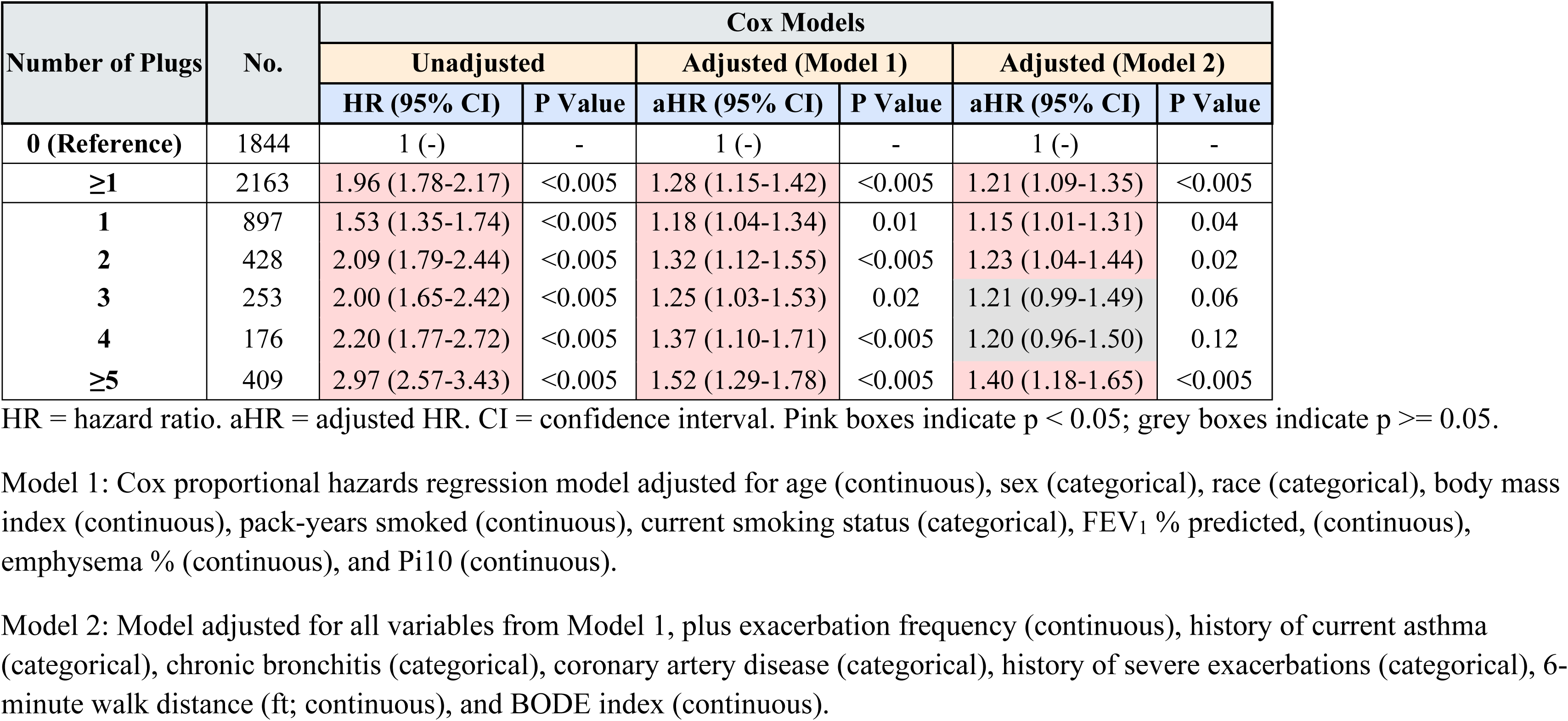
Results of Cox mortality analysis for GOLD 1–4 cases.

Fine–Gray competing risk models were used to evaluate cause-specific mortality (**Figure 5**). Compared with participants without plugs, plug presence was associated with increased respiratory mortality across GOLD stages, with the largest relative risk observed in GOLD 0 (aHR, 4.23; 95% CI, 1.29–13.90; P = 0.018), and significant associations in GOLD 1–2 (aHR, 1.62; 95% CI, 1.07–2.44; P = 0.022) and GOLD 3–4 (aHR, 1.30; 95% CI, 1.04–1.62; P = 0.021). In PRISm, the association was elevated but not statistically significant (aHR, 2.31; 95% CI, 0.69–7.76; P = 0.18). Cardiovascular mortality was significantly associated with plug presence only in GOLD 1–2 (aHR, 1.54; 95% CI, 1.03–2.30; P = 0.037), whereas cancer mortality was not significantly associated with plugs in any subgroup.

**Figure 5.**
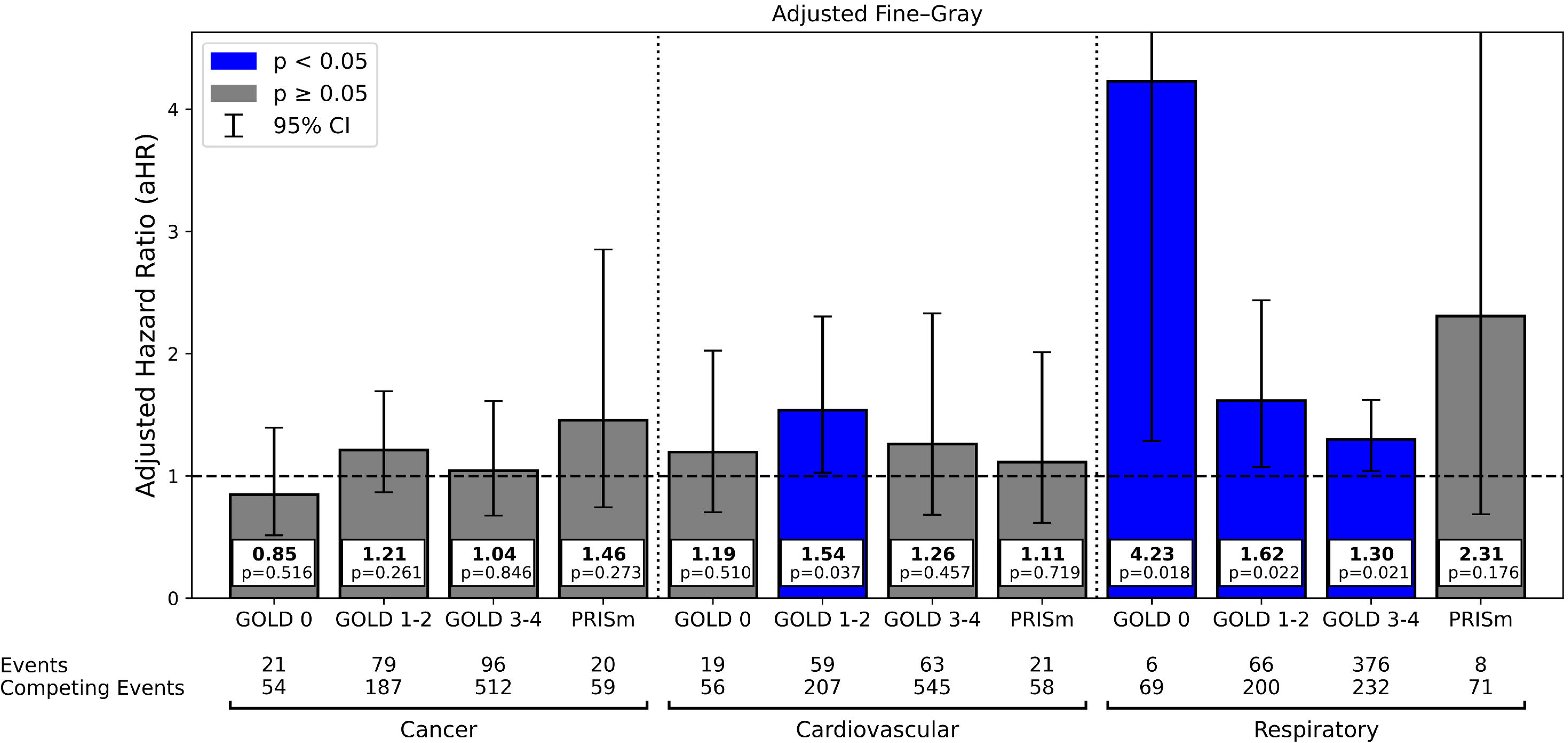
Fine–Gray sub distribution hazards analysis for cause-specific mortality by GOLD subgroup. Adjusted hazard ratios (aHRs) compare cases with plugs to those without. Results adjusted for all Model 1 adjustments— age (continuous), sex (categorical), race (categorical), body mass index (continuous), pack-years smoked (continuous), current smoking status (categorical), FEV1 % predicted, (continuous), emphysema % (continuous), and Pi10 (continuous) —and comorbidities relevant to specific causes of death (all categorical): coronary artery disease, lung, breast, prostate, colon, and bladder cancer, congestive heart failure, stroke, diabetes, and high blood pressure.

## Discussion

In this large, well-characterized cohort of tobacco-exposed individuals, we applied an automated quantitative CT framework to identify and quantify airway mucus plugging. Increased mucus plug burden was associated with airflow limitation, CT-defined air trapping, greater symptom burden, and reduced exercise capacity. Importantly, higher burden was independently associated with exacerbation risk, all-cause mortality, and respiratory mortality after adjustment for established clinical and imaging covariates. These findings extend prior visual CT studies by providing scalable, objective quantification and support airway mucus plugging as a clinically relevant airway phenotype that contributes to disease heterogeneity in COPD.

### Challenges in Defining and Identifying Mucus Plugging

Although airway mucus plugging is increasingly recognized as clinically relevant, its CT definition remains variable across studies. A standardized consensus definition has not been established, and most investigations rely on visually defined, segment-based scoring systems that classify plugs as present or absent (21). These approaches are reader dependent, demonstrate only moderate interobserver agreement (3), and typically require complete luminal occlusion, potentially underestimating partial or low-density obstruction.

Manual annotation of mucus plugs requires specialized expertise and is time-intensive, limiting scalability for large cohort studies and routine clinical application. Prior studies have reported substantial inter-reader variability in visual mucus plug scoring, underscoring the need for more reproducible approaches. These constraints pose practical barriers to consistent phenotyping of airway obstruction across populations.

Our automated framework approaches this challenge from an anatomic perspective by identifying airway segmentation discontinuities consistent with luminal obstruction rather than relying on explicit plug-level annotations. This strategy reduces dependence on manual ground-truth labeling and avoids the need for synthetic plug generation during model development (15). By leveraging lumen-based airway modeling, the approach provides an objective and scalable measure of mucus plug burden while allowing flexibility as definitions evolve. Automated quantitative methods such as this may facilitate large-scale research applications and support future integration into airway-focused imaging analyses.

### Clinical and Translational Implications of Automated Mucus Plug Quantification

Although many of the clinical associations observed in this study have been reported previously using visual CT scoring, a major contribution of this work is the development and validation of an automated AI-based framework for quantitative assessment of airway mucus plugging. Widespread implementation of mucus plug phenotyping has been limited by the need for expert visual review, which is subjective, time-consuming, and difficult to scale across large cohorts or routine clinical practice.

Our approach addresses these limitations by providing objective and reproducible quantification without extensive plug-level annotation. Importantly, the automated measurements reproduced many of the established associations between mucus plugging and airflow limitation, exacerbations, and mortality. The concordance between automated and visual approaches suggests that clinically meaningful information can be extracted reliably without manual scoring.

As quantitative CT analyses become increasingly integrated into pulmonary imaging, automated mucus plug quantification may facilitate broader incorporation of this airway phenotype into research and clinical workflows. Routine availability of mucus plug measurements could enhance patient phenotyping, improve risk stratification, and support future studies evaluating airway-targeted therapies and interventions aimed at mucus clearance.

### Interpretation of Findings

Across the COPDGene cohort, increasing mucus plug burden was associated with progressive physiologic impairment. FEV₁ % predicted declined steadily with higher burden, and PRM-defined air trapping increased in parallel, consistent with greater small airway dysfunction. Symptom burden and functional limitation followed a similar pattern, with worse SGRQ scores and shorter 6-minute walk distance observed at higher plug levels. Although these relationships were most pronounced between 0 and 2 plugs, additional increases in burden were associated with more modest incremental changes in symptom and exercise measures.

These findings are consistent with prior visual CT studies. Dunican et al. (2) reported that visually scored mucus plugs in SPIROMICS were associated with lower FEV₁, greater exacerbation risk, reduced exercise capacity, and more advanced GOLD stage—findings that parallel those observed in COPDGene. The concordance between cohorts using distinct assessment methods supports the robustness of mucus plugging as a marker of airway disease severity.

Collectively, the data suggest that even modest increases in mucus plug burden correspond to clinically meaningful impairment and adverse outcomes. The observed attenuation of symptom and exercise associations at higher burden levels may reflect ceiling effects in functional measures rather than absence of additional physiologic impact.

### Association with Exacerbations

Mucus plug presence was independently associated with both exacerbation frequency and severe exacerbations after multivariable adjustment. These findings are consistent with the concept that airway luminal obstruction may contribute to ventilation heterogeneity, impaired secretion clearance, and susceptibility to infectious or inflammatory events. Notably, the association persisted after adjustment for airflow limitation, emphysema burden, airway wall thickness, and prior exacerbation history, suggesting that mucus plugging captures a dimension of airway pathology not fully reflected by conventional physiologic or structural measures.

Our results align with prior visual CT studies demonstrating associations between mucus plugging and exacerbation risk in COPD. The reproducibility of these relationships using automated quantitative assessment further supports mucus plugging as a clinically meaningful airway phenotype. While the observational nature of this analysis precludes causal inference, the consistency of associations across physiologic impairment, exacerbations, and mortality highlights the potential relevance of airway mucus obstruction in disease progression.

### Association with Mortality

Higher mucus plug burden was associated with increased all-cause mortality, and this relationship persisted after multivariable adjustment for demographic, clinical, and imaging covariates. Although effect estimates varied modestly across burden categories, the presence of mucus plugs was consistently associated with greater mortality risk compared with plug-free cases across unadjusted and adjusted models.

These findings are consistent with prior analyses in COPDGene. Diaz et al. (11) demonstrated that visually scored mucus plug burden was independently associated with increased mortality, and van der Veer et al. reported comparable associations using an automated approach with similar covariate adjustment. The concordance across visual and automated methods supports the robustness of mucus plugging as a prognostic marker.

In cause-specific analyses, mucus plug presence was associated with increased respiratory mortality across GOLD stages, with the largest relative association observed in GOLD 0. A significant association with cardiovascular mortality was observed in GOLD 1–2. Although prior work has reported stage-dependent variation in cause-specific associations, the overall pattern across studies suggests that greater mucus plug burden is linked to increased respiratory mortality risk.

### Limitations and Future Directions

Several limitations warrant consideration. First, the absence of a standardized consensus definition of mucus plugging complicates comparison across studies and imaging approaches. Although our automated framework reduces reliance on manual labeling, external validation across diverse scanners, acquisition protocols, and populations remains necessary. Plug detection was based on inspiratory CT alone, and dynamic or expiratory changes in airway obstruction were not evaluated.

Second, this analysis was observational and plug burden was assessed at a single time point, precluding inference regarding causality or temporal variability. Although multivariable adjustment was extensive, residual confounding—including treatment patterns and comorbid conditions—cannot be excluded.

Third, automated identification of airway discontinuities depends on segmentation performance and CT spatial resolution. Distal or partially occluding mucus plugs below imaging resolution may be under detected, and airway distortion in advanced disease could influence segmentation accuracy despite robust validation.

Future studies should evaluate model generalizability across additional cohorts and airway diseases and incorporate longitudinal imaging to determine whether mucus plug burden is stable, progressive, or reversible over time. Assessing whether changes in plug burden correspond to disease progression, exacerbation risk, or therapeutic response will be important for clarifying clinical relevance. Integration with regional physiologic measures and biologic markers of mucus dysfunction may further elucidate mechanistic pathways and refine airway phenotyping.

## Conclusions

In this large, well-characterized cohort of tobacco-exposed individuals, automated AI-based quantitative CT assessment of airway mucus plugging identified a reproducible airway phenotype associated with airflow limitation, greater symptom burden, increased exacerbation risk, and higher mortality. These associations persisted after adjustment for established clinical and imaging markers of disease severity, and plug presence was independently associated with increased respiratory mortality across GOLD stages. By providing objective, scalable quantification without reliance on manual visual scoring, this approach extends prior observations and enables consistent phenotyping of airway obstruction at population scale. Collectively, these findings support airway mucus plugging as a clinically relevant dimension of COPD heterogeneity and highlight the potential of automated quantitative assessment to improve risk stratification and facilitate future longitudinal studies and therapeutic investigations targeting airway mucus dysfunction.

## Data Availability

The datasets presented in this study are not readily available because they are part of NIH sponsored clinical trials and require signing of a data use agreement. For instructions on how to access COPDGene
data visit https://www.copdgene.org/phase-1-study-documents.htm.

## Acknowledgements

This work was supported by Department of Defense grant HT94252310698 and National Heart, Blood, and Lung Institute grants R01HL162661, U01 HL089897 and U01 HL089856 and by National Institutes of Health contract 75N92023D00011. The COPDGene study (NCT00608764) has also been supported by the COPD Foundation through contributions made to an Industry Advisory Committee that has included AstraZeneca, Bayer Pharmaceuticals, Boehringer-Ingelheim, Genentech, GlaxoSmithKline, Novartis, Pfizer, and Sunovion.

