## Supplementary_Material for "Artificial Intelligence-Based Detection of Airway Mucus Plugs on CT and Associations With Clinical Outcomes in COPDGene"

### I. Supplemental Methods

#### A. Parametric Detection of Airway Discontinuities

##### i. *Airway Discontinuity Detection and Candidate Ranking*

Following airway segmentation, airway centerlines were skeletonized and represented as a graph of connected airway branches. Potential airway discontinuities were defined as disconnected components within the airway skeleton and were identified as candidate anomalies for further evaluation.

Two classes of candidate anomalies were considered:

1. *Endpoint–endpoint connections*, in which disconnected terminal endpoints from separate components may represent interruption of a continuous airway branch.
2. *Endpoint–nonendpoint connections*, in which a terminal endpoint may connect to a nonterminal point on another component, enabling identification of plugs occurring near airway bifurcations.

Candidate anomalies were iteratively incorporated into the airway tree using a parametric loss function designed to favor anatomically plausible reconnections while excluding unlikely discontinuities.

##### ii. *Geometric Constraints*

For each candidate anomaly, an airway direction vector was defined from neighboring skeleton points adjacent to the candidate endpoint. Angular deviation introduced by a proposed reconnection was quantified using

$$\theta = \cos^{-1}\left(\frac{u \cdot v}{|| u |||| v ||}\right)$$

where $u$ and $v$ denote airway direction vectors.

Small, disconnected components (<5 voxels) were excluded, as these frequently represented false-positive fragments with insufficient topology for reliable direction estimation.

##### iii. *Candidate Ranking*

Candidate anomalies were ranked using loss functions based on Euclidean connection distance (*d*) and angular deviation ($\theta$).

For endpoint–endpoint candidates:

$$L(d, \theta_1, \theta_2) = \begin{cases} \infty & d > 50 \\ \infty & \theta_1 > 65^\circ \text{ or } \theta_2 > 65^\circ \\ d + 0.1(\theta_1 + \theta_2) & \text{otherwise} \end{cases}$$

For endpoint–nonendpoint candidates:

$$L(d, \theta) = \begin{cases} \infty & d > 50 \\ \infty & \theta > 40^\circ \\ d + 0.2\theta & \text{otherwise} \end{cases}$$

Candidates exceeding predefined distance or angular thresholds were excluded from consideration. Remaining candidates were ranked by loss, with lower values indicating greater anatomic plausibility.

**iv.** *Candidate Discontinuity Output*

Retained candidate discontinuities represented localized airway breaks potentially corresponding either to segmentation incompleteness or true luminal occlusion. These candidate regions were then passed to the downstream convolutional neural network classifier for discrimination of mucus plugs from unobstructed airway and parenchymal regions.

#### B. Classification of Candidate Airway Obstructions

**i.** *Classification of Candidate Discontinuities*

Candidate airway discontinuities identified in Step 2 were classified as mucus plug, unobstructed airway, or parenchyma using a lightweight one-dimensional convolutional neural network (CNN). For each candidate discontinuity, a one-dimensional voxel-intensity patch of 30 voxels in length was sampled along the predicted airway centerline, spanning the discontinuity and adjacent airway segments. This representation preserves local attenuation structure and transition patterns relevant to airway obstruction.

**ii.** *Model-Based Construction of the Mucus Plug Class*

Training samples for unobstructed airway and parenchyma classes were obtained from visually confirmed normal airway and lung regions.

Because large-scale plug-level annotations are limited, the mucus plug class was constructed using a weakly supervised, model-based strategy. A small set of visually confirmed, fully occlusive mucus plugs was used to characterize attenuation profiles associated with luminal obstruction. These empirical attenuation values were modeled using kernel density estimation

(KDE) with an Epanechnikov kernel to estimate the probability density of voxel intensities within obstructed airways:

$$\hat{f}(x) = \frac{1}{nh}\sum_{i=1}^{n} K\left(\frac{x - x_i}{h}\right)$$

where $x_i$ denotes attenuation samples from confirmed plug regions, $h$is the bandwidth parameter, and $n$is the number of sampled voxels.

The Epanechnikov kernel was defined as:

$$K(u) = \begin{cases} \frac{3}{4}(1 - u^2), & |\, u \,| \leq 1 \\ 0, & |\, u \,| > 1 \end{cases}$$

This kernel provides compact support and efficient localized density estimation, enabling realistic modeling of attenuation variability within mucus plugs.

The estimated density function was used to generate representative plug attenuation profiles, with additional uniform perturbations incorporated to simulate variability in plug extent and morphology along airway segments. This approach enabled construction of realistic mucus plug training examples without extensive manual annotation or reliance on purely synthetic plug generation.

**iii.** *CNN Architecture and Classification*

A lightweight one-dimensional CNN was trained to classify candidate discontinuities using the extracted centerline attenuation patches.

The network architecture consisted of two sequential one-dimensional convolutional layers with rectified linear unit (ReLU) activations:

- The first convolutional layer learned 16 filters to capture local attenuation patterns.
- The second convolutional layer learned 32 filters to extract higher-order feature representations of airway continuity and obstruction.

The resulting 32 feature maps, each of length 30, were flattened and concatenated into a 960 × 1 feature vector (32 × 30). This feature vector was passed through a fully connected layer to generate class probabilities for three categories: mucus plug, unobstructed airway, and parenchyma.

Model optimization was performed using categorical cross-entropy loss:

$$L = -\sum_{c=1}^{C} y_c \log(p_c)$$

where $y_c$ denotes class labels and $p_c$ predicted class probabilities.

Candidate discontinuities classified as mucus plugs were aggregated to compute participant-level mucus plug burden, defined as total plug count.

#### C. Network Training and Implementation Details

The nnU-Net–based airway segmentation model was trained from scratch using the standard nnU-Net training framework. Training was performed for 1,000 epochs, with each epoch consisting of 250 mini-batches. Network parameters were optimized using stochastic gradient descent with Nesterov momentum (μ = 0.99) and an initial learning rate of 0.01. Instance normalization was applied before each Leaky ReLU activation to improve training stability and facilitate convergence. Following the nnU-Net framework, global normalization statistics were updated using instance-specific estimates during training, as previously described (1). Standard nnU-Net data augmentation strategies, including geometric and intensity transformations, were used to improve model robustness and generalizability.

The lightweight one-dimensional CNN classifier was likewise trained from scratch using randomly initialized weights. Model optimization was performed using the Adam optimizer with an initial learning rate of 0.01 and a batch size of 50. Training was conducted for 100 epochs using categorical cross-entropy loss. To reduce overfitting and improve generalization, $L_2$ weight decay regularization ($\lambda = 1 \times 10^{-5}$) and a dropout rate of 0.3 were applied during training. Training samples were constructed to ensure balanced representation of mucus plug, unobstructed airway, and parenchyma classes.

Both the nnU-Net airway segmentation model and the lightweight CNN classifier were implemented in Python using the PyTorch deep learning framework and trained on an NVIDIA Tesla V100 GPU with 16 GB of memory. Model development and evaluation were performed using independent training and testing datasets, and all reported performance metrics were obtained on previously unseen test data.

#### D. Mucus Plug Burden Metrics

Three participant-level measures of mucus plug burden were derived from the detected plug locations.

**i.** *Mucus Plug Count*

Mucus plug count was defined as the total number of detected airway obstructions within a participant.

**ii.** *Affected Airways Score*

To quantify both the local extent of obstruction and the amount of airway tree affected by each plug, an affected airways score was calculated as:

$$\text{Affected Airways Score} = \sum_{p=1}^{N} \left(\frac{l_p}{l_s^p}\right)\left(\frac{V_d^p}{V_a}\right)$$

where,

$l_p$ = the length of the plug denoting the longitudinal extent of the obstruction within the affected airway segment

$l_s^p$ = the length of the airway segment in which the plug $p$ resides

$V_d^p$ = the volume of airways downstream to the plug $p$ denoting the volume of airway branches distal to the obstruction

$V_a$ = is the total airway segmented volume for the participant.

This metric assigns greater weight to plugs occupying a larger proportion of an airway segment and occurring in locations supplying larger portions of the airway tree.

**iii.** *Weibel Sum*

A generation-weighted burden metric was also calculated using the Weibel generation associated with each detected plug:

$$\text{Weibel(Plug)} = \max\left(2^{(23-\text{generation})} - 1, 0\right);$$

$$\text{Weibel Sum} = \sum_{\text{plugs}} \text{Weibel(plug)}$$

This metric assigns progressively greater weight to plugs occurring in proximal airways because obstruction at earlier airway generations may affect a larger volume of distal lung.

**iv.** *Selection of Primary Burden Metric*

Associations between all three burden metrics and physiologic and clinical outcomes were evaluated (**Supplemental Figure 4**). Because the three measures yielded highly similar relationships across outcomes, mucus plug count was selected as the primary burden metric owing to its simplicity, interpretability, and direct clinical relevance. All subsequent analyses therefore use total mucus plug count as the primary measure of mucus plug burden.

**Supplementray Figure 1**

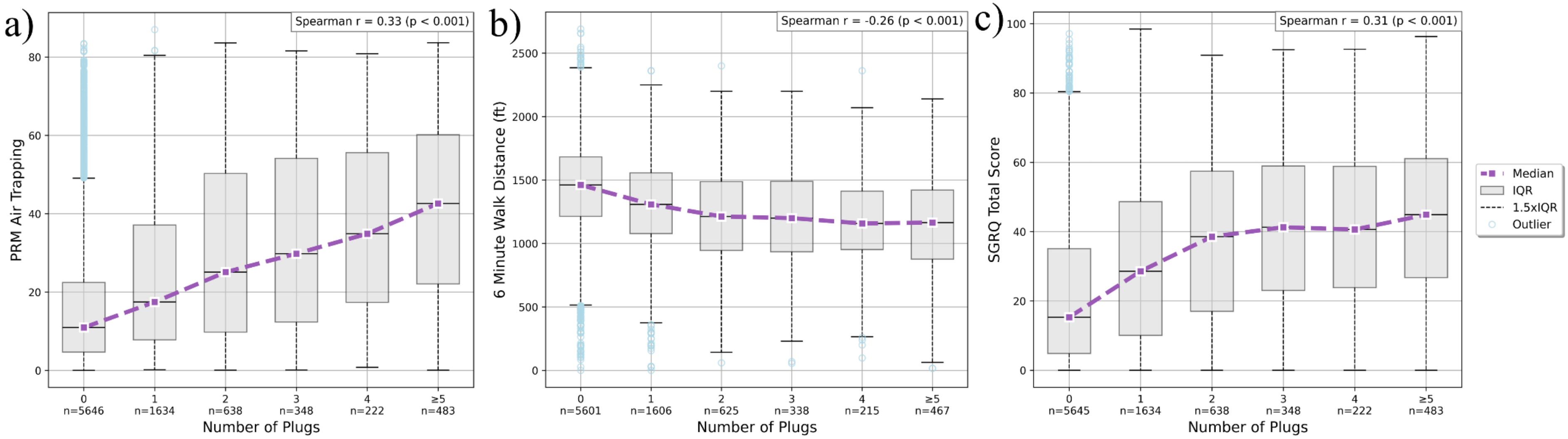


Figure (a) shows the relationship between number of plugs and PRM air trapping; figure (b) shows the relationship between number of plugs and 6-minute walk distance; figure (c) shows the relationship between number of plugs and SGRQ total score. The x-axis shows the number of plugs (0, 1, 2, 3, 4, ≥5), and the y-axis shows median value for each plug count, with interquartile range displayed. Outliers are shown in blue. Spearman correlation coefficient (r) and p-value are displayed on the top right of the plot. For display purposes, participants with ≥5 plugs were grouped; correlation analyses were conducted using continuous burden values.

**Supplementary Figure 2**

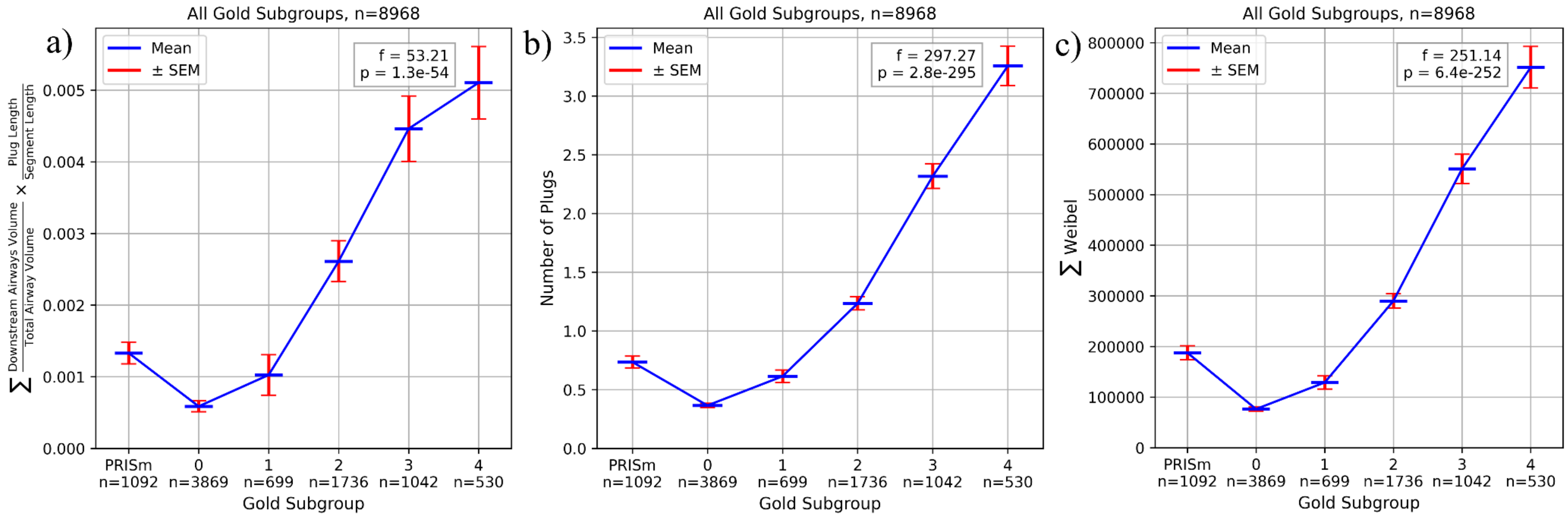


A plot of (a) mean affected airways score, (b) mean number of plugs and (c) mean Weibel sum by GOLD subgroup.

**Supplementary Figure 3**

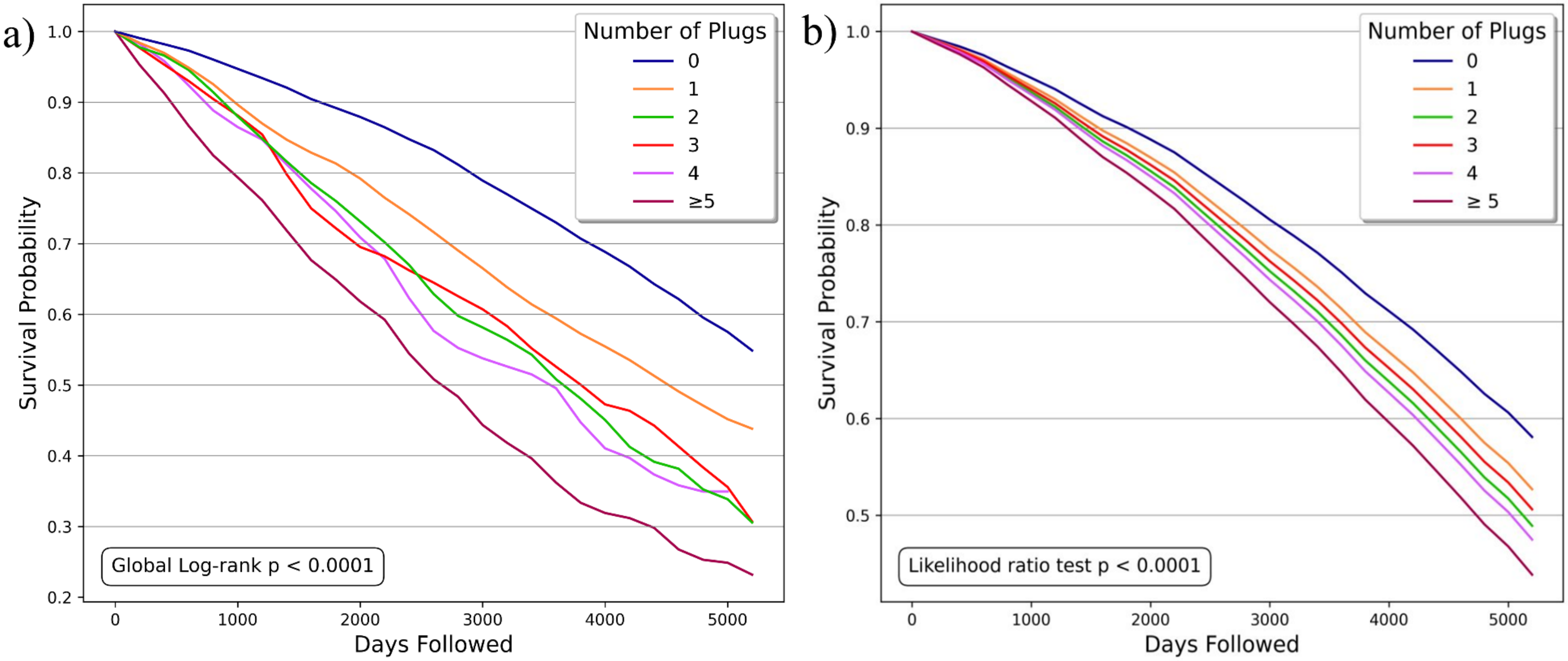


Results of survival analysis for GOLD 1–4 cases by number of plugs. Cases with 0, 1, 2, 3, 4, and ≥5 plugs at baseline are analyzed separately. Figure (a) shows unadjusted Kaplan–Meier analysis. Figure (b) shows survival probability predictions over time from an adjusted Cox proportional hazards model with adjustment for age, sex, race, body mass index, pack-years smoked, current smoking status, $FEV_1$, emphysema %, and Pi10.

**Supplementary Figure 4**

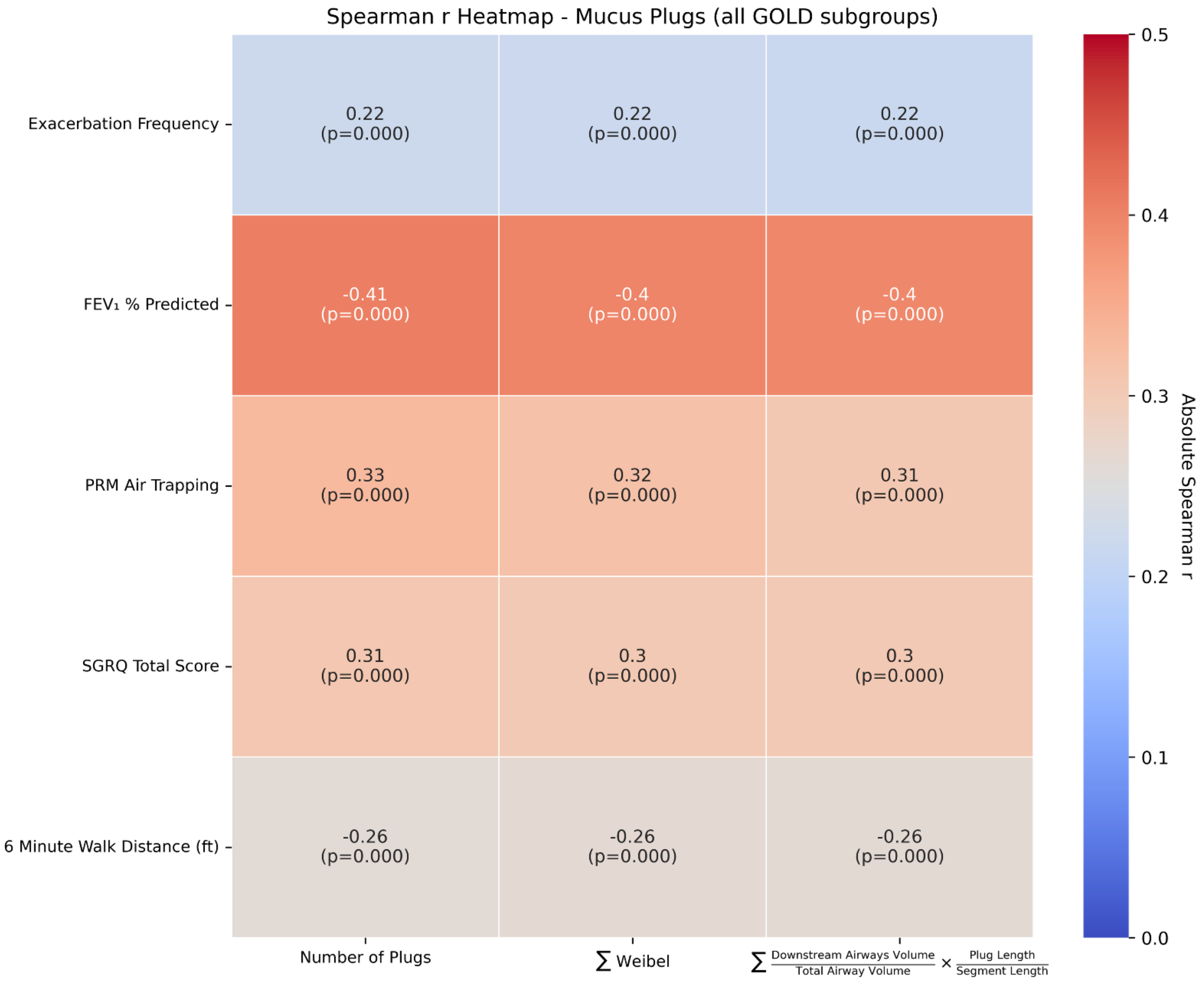


Spearman correlation results separately comparing number of plugs, Weibel sum, and affected airways score to exacerbation frequency, $FEV_1$ % predicted, PRM Air Trapping, SGRQ total score, and 6-minute walk distance.
